# Emergence of multiple SARS-CoV-2 antibody escape variants in an immunocompromised host undergoing convalescent plasma treatment

**DOI:** 10.1101/2021.04.08.21254791

**Authors:** Liang Chen, Michael C Zody, Jose R Mediavilla, Marcus H Cunningham, Kaelea Composto, Kar Fai Chow, Milena Kordalewska, André Corvelo, Dayna M Oschwald, Samantha Fennessey, Marygrace Zetkulic, Sophia Dar, Yael Kramer, Barun Mathema, Tom Maniatis, David S Perlin, Barry N Kreiswirth

## Abstract

SARS-CoV-2 Variants of Concerns (VOC), e.g., B.1.351 (20H/501Y.V2) and P1 (20J/501Y.V3), harboring N-terminal domain (NTD) or the receptor-binding domain (RBD) (e.g., E484K) mutations, exhibit reduced in vitro susceptibility to convalescent serum, commercial antibody cocktails, and vaccine neutralization, and have been associated with reinfection. The accumulation of these mutations could be the consequence of intra-host viral evolution due to prolonged infection in immunocompromised hosts. In this study, we document the microevolution of SARS-CoV-2 recovered from sequential tracheal aspirates from an immunosuppressed patient on tacrolimus, steroids and convalescent plasma therapy, and identify the emergence of multiple NTD and RBD mutations associated with reduced antibody neutralization as early as three weeks after infection. SARS-CoV-2 genomes from the first swab (Day 0) and three tracheal aspirates (Day 7, 21 and 27) were compared at the sequence level. We identified five different S protein mutations at the NTD or RBD regions from the second tracheal aspirate sample (21 Day). The S:Q493R substitution and S:243-244LA deletion had ∼70% frequency, while ORF1a:A138T, S:141-144LGVY deletion, S:E484K and S:Q493K substitutions demonstrated ∼30%, ∼30%, ∼20% and ∼10% mutation frequency, respectively. However, the third tracheal aspirate sample collected one week later (Day 27) was predominated by the haplotype of ORF1a:A138T, S:141-144LGVY deletion and S:E484K (> 95% mutation frequency). Notably, S protein deletions (141-144LGVY and 243-244LA deletions in NTD region) and substitutions (Q493K/R and E484K in the RBD region) previously showed reduced susceptibly to monoclonal antibody or convalescent plasma. The observation supports the hypothesis that VOCs can independently arise and that immunocompromised patients on convalescent plasma therapy are potential breeding grounds for immune-escape mutants.

**Competing Interest Statement:** The authors have declared no competing interest.

**Funding Statement:** The study was in part supported by Center for Discovery and Innovation and Hackensack Meridian Health Foundation.

## Introduction

After a year of the COVID-19 pandemic, with over 100 million global cases and 2.80 million deaths, the world is now focused on the biological consequences of the distribution of vaccines and the spread of “Variants of Concerns” (VOC). Three SAR-CoV-2 VOCs, i.e., B.1.1.7 (20I/501Y.V1), B.1.351 (20H/501Y.V2) and P1 (20J/501Y.V3) carrying the spike protein N501Y mutation emerged in the UK, South Africa, Brazil and Japan^1^, and have been associated with high transmissibility due to the increased affinity to the ACE receptor. In each of these viruses, the spike protein contains clustered mutations in the N-terminal domain (NTD) and the receptor-binding domain (RBD) (e.g., E484K) regions. Some VOCs carrying these mutations show reduced *in vitro* susceptibility to convalescent serum, commercial antibody cocktails, and vaccine neutralization, and have been associated with reinfection^2,3^. The accumulation of these mutations is assumed to be the consequence of intra-host viral evolution due to prolonged infection in immunocompromised hosts^4,5^. A recent NEJM report from Choi et al.^4^ described the emergence of antibody escape mutations from an immunocompromised patient 75 days after infection. Here, we document the microevolution of SARS-CoV-2 recovered from sequential tracheal aspirates from an immunosuppressed patient on tacrolimus, steroids and convalescent plasma therapy, and identify the emergence of multiple NTD and RBD mutations associated with reduced antibody neutralization as early as three weeks after infection.

## Materials and Methods

### SARS-CoV-2 detection

Total nucleic acid (TNA) from nasopharyngeal swabs was extracted by the MagNAPure 24 system (Roche Life Science) and Viral RNA from tracheal aspirates was extracted using QIAamp Viral RNA Mini Kit (Qiagen), following the manufacturer’s instructions. SARS-CoV-2 detection was performed using the CDI-enhanced COVID-19 test^6^, targeting SARS-CoV-2 E an N2 genes. The test was approved for use on March 12, 2020 under FDA Emergency Use Authorization for COVID-19 and has a limit of detection less than 20 viral genome copies per reaction. A specimen is considered positive if the gene target has a cycle threshold (Ct) value < 40.

### SARS□CoV□2 viral sequencing and genomic analysis

SARS-CoV-2 targeted assay libraries were prepared using the AmpliSeq Library Plus and cDNA Synthesis for Illumina kits (Illumina) in accordance with manufacturer’s recommendations. Briefly, 20ng of RNA was reverse transcribed followed by amplification of cDNA targets using the Illumina SARS-CoV-2 research panel (Illumina). The amplicons were then partially digested, ligated to AmpliSeq CD Indexes, and then amplified using 18 cycles of PCR. Libraries were quantified using fluorescent-based assays including PicoGreen (Life Technologies), Qubit Fluorometer (Invitrogen), and Fragment Analyzer (Advanced Analytics). Final libraries were sequenced on a NovaSeq 6000 sequencer (v1 chemistry) with 2×150bp.

Short read-data were filtered and processed prior to alignment. Read pairs that did not contain a single 19bp seed k-mer in common with the SARS-CoV-2 genome reference (NC_045512.2) were discarded. Adapter sequences and low quality (Q < 20) bases were trimmed from the remaining reads, using Cutadapt v2.101^7^. Processed reads were then mapped to the SARS-CoV-2 genome reference using BWA-MEM v0.7.172^8^ and only read pairs with at least one alignment spanning a minimum of 42 bp in the reference and starting before position 29,862 (to exclude polyadenine-only alignments) were kept. Genome sequences were determined by alignment pileup consensus calling with a minimum support of 5 reads using Samtools v1.11 and bcftools v1.11 ^9^. SNP and InDels were called using FreeBayes v1.3.5 (https://github.com/freebayes), followed by annotation using SnpEff v4.5^10^. A minimum variant calling frequency was set to be 5% to identify within host variations.

The resulting SARS-CoV-2 viral genome sequences were uploaded to Nextclade server (https://clades.nextstrain.org/) to assign Nextstrain clades^11^. SARS-CoV-2 lineage was determined using Pangolin v2.3.0 (https://github.com/cov-lineages/pangolin) and GISAID clade is determined based upon the clade specific marker variants from https://www.gisaid.org^12^. In addition, 2,282 SARS-CoV-2 genomes with E484K mutation were downloaded from GISAID database^12^ (date as 2/12/2021), and the genomes with less than 1% ambiguous nucleotides (Ns) and > 28,900 bp were aligned using MAFFT v7.475 ^12^ using default setting. A maximum likelihood phylogenetic tree was constructed using IQ-TREE v2.1.2^13^ with automatic model selection and 1000-bootstrap replicates. The resulting tree was annotated using ITOL v5^14^.

### Informed consent

Informed consent was obtained from this patient and the study was approved by Hackensack Meridian Health Institutional Review Board (IRB) under protocol Pro2018-1022.

## Results

### Case description

A male in early 50s presented to a Northern Jersey hospital with fever, productive cough, generalized myalgias, and progressive shortness of breath for 4 days (**Fig1A**). He had history of deceased donor kidney transplant for end-stage renal disease (ESRD) secondary to HTN, complicated by cellular graft rejection and recurrent collapsing focal segmental glomerulosclerosis. On physical examination, the patient had fever of temperature 102.3F, O2 Saturation 90% on 100% non-rebreather. His examination was significant for tachypnea, but otherwise unremarkable. He was admitted under the suspicion of COVID-19 pneumonia. His medications were significant for his immunosuppressive regime of mycophenolic acid, prednisone, and tacrolimus along with multiple anti-hypertensive medications.

**Fig1.**
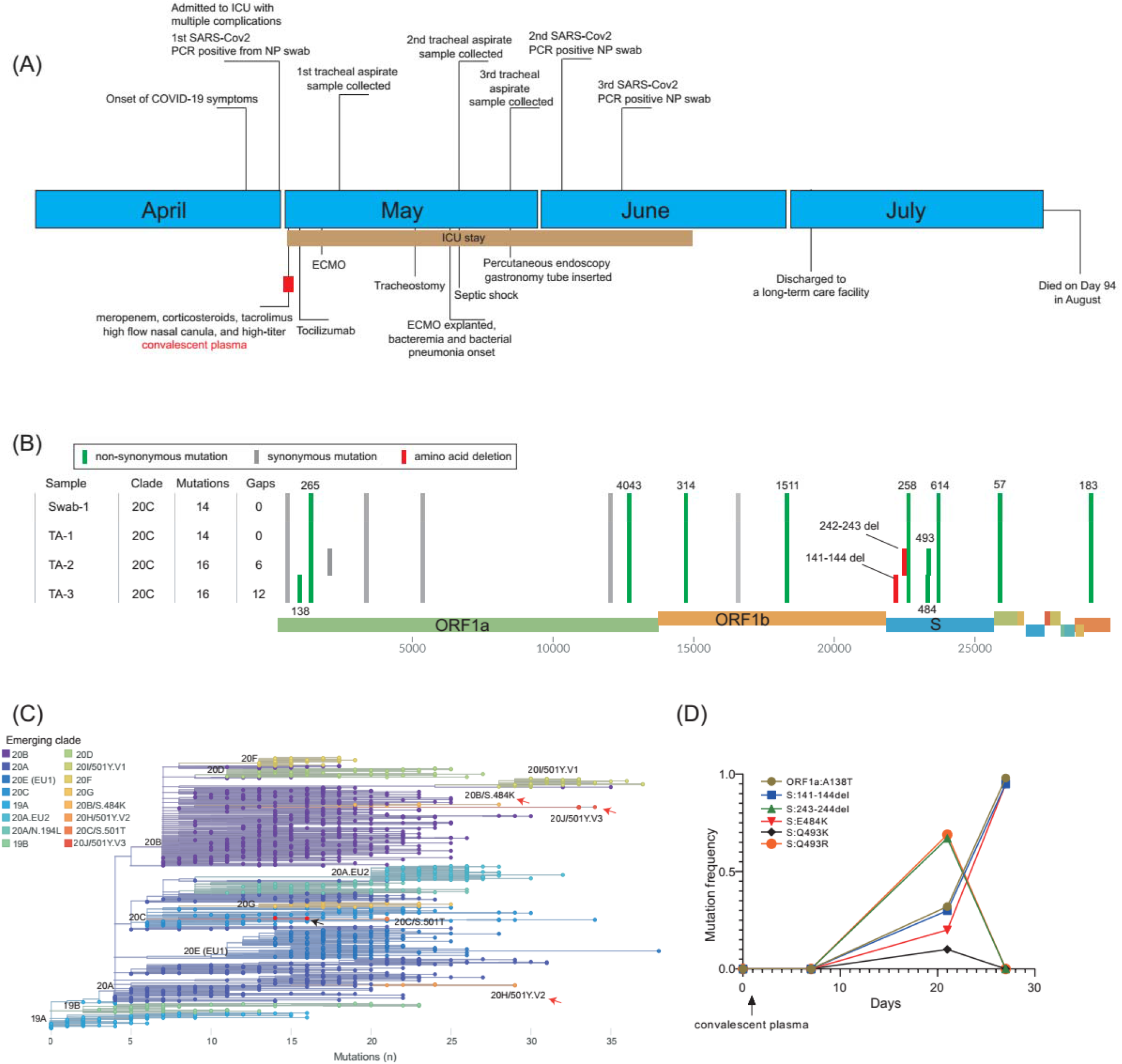
Clinical and genomic characterization of SARS-CoV-2 variations in an immunocompromised patient. (**A)**, Clinical timeline of events of the immunocompromised patient. (**B**), SARS-CoV-2 genotypes of the major haplotypes from the swab (swab-1) and tracheal aspirate samples (TA-1, day 7; TA-2, day 21 and TA-3, day 27). (**C**), The phylogenetic clades of SARS-CoV-2 variants. The tree was generated using Nextclade (clades.nextstrain.org). The nodes are highlighted by the Nextstrain clades. The clades associated with E484K mutations are denoted by red arrows, while the B.1.369 genomes described in this study are illustrated by a black arrow. (**D**), The mutation frequency changes among the swab and tracheal aspirate samples over time.

COVID-19 was confirmed to be positive by RT-PCR (Day 0). Chest X-Ray (CXR) revealed dense infiltrates bilaterally reflective of his viral pneumonia. Given his multiple comorbidities, immunosuppressed status, and work of breathing, he was admitted to the ICU for high flow and awake proning and was started on broad spectrum meropenem treatment. His anti-hypertensives were discontinued due to his normotension, and his immunosuppressive regime was continued except for mycophenolate given the likelihood of serious infection.

He was treated with high-titer convalescent plasma (Day 1) and tocilizumab (Day 2). Due to his worsening respiratory status, the patient was intubated (Day 2). Antibiotics were switched to vancomycin and piperacillin-tazobactam and then discontinued as the patient was afebrile (Day 3). The patient was found to have bilateral deep venous thrombosis and was started on therapeutic heparin (Day 3). Due to worsening hypoxic respiratory failure despite complete support from mechanical ventilation, the patient was subsequently cannulated and placed on veno-venous extra-corporeal oxygenation (ECMO) (Day 5). He went into rapid atrial flutter as well and was started on intravenous amiodarone (Day 5). His renal failure attributed to multiple factors such as his tacrolimus, COVID-19 injury, and hypotension slowly began to improve. Oxygenation began to improve and stabilize, leading to tracheostomy (Day 16) and ECMO explantation (Day 20). The patient, however, became febrile and septic with *Enterococcus* bacteremia and *Proteus mirabilis* pneumonia and was restarted on vancomycin and piperacillin-tazobactam (Day 20). He subsequently developed septic shock and was started on vasopressors (Day 21). Following the antibiogram, antibiotics were de-escalated to ampicillin (Day 21) and continued for a 7-day course. The septic shock resolved, and the patient was re-started on his anti-hypertensives once his blood pressure began to remain stable. His course continued to be complicated by periodic desaturations and wide and narrow complex tachycardia, anemia, and thrombocytopenia. He slowly improved permitting ventilation and sedation weaning. As his dysphagia was unresolved during his recovery, percutaneous endoscopic gastrostomy (PEG) tube was placed (Day 28) to improve nutritional status. He was transferred to the step-down unit as he continued to recover (Day 49) and was discharged to a long-term care facility (Day 64) requiring ventilatory support only at night. Unexpectedly, the patient expired presumably due to hypoxic respiratory failure secondary to his COVID-19 pneumonia (Day 94).

### Genomic analysis

SARS-CoV-2 positive qRT-PCR results (**Table 1**) were obtained from three nasopharyngeal swab samples (on Day 0, 34 and 41) and three tracheal aspirates (on Day 7, 21 and 27); the first swab and the three tracheal aspirates were available for viral genome sequencing (**Fig1A**). The genotype of the initial swab and tracheal aspirate (Day 7) were identical. The genomes of these two samples harbored 14 mutations (versus Wuhan-Hu-1), and were assigned as Nextstrain clade 20C, Pangolin lineage B.1.369 and GISAID clade GH, distinct from the three 501Y VOCs (**Fig1B, C**). The second tracheal aspirate specimen (from Day 21) showed five different S protein mutations at the NTD or RBD regions. The S protein Q493R substitution and 243-244LA deletion had ∼70% frequency, while ORF1a A138T, S protein 141-144LGVY deletion, E484K and Q493K substitutions demonstrated ∼30%, ∼30%, ∼20% and ∼10% mutation frequency, respectively (**Fig1D**). However, the third tracheal aspirate sample collected one week later (Day 27) was predominated by the haplotype of ORF1a:A138T, S:141-144LGVY deletion and S:E484K (> 95% mutation frequency) (**Fig1D**).

**Table 1.** Cycle-threshold values of SARS-CoV-2 samples.

| Sample | Ct for N2 target |
| --- | --- |
| Swab-1 (Day 0) | 24.34 |
| TA-1 (Day 7) | 20.43 |
| TA-2 (Day 21) | 15.39 |
| TA-3 (Day 27) | 24.65 |
| Swab-2 (Day 34) | 37.14 |
| Swab-3 (Day 41) | 32.39 |

The 141-144LGVY and 243-244LA deletions are located in the recently described “Recurrent Deletion Region” (RDR) 2 and 4^15^, respectively, within the NTD of the spike protein. The appearance of deletions in the RDR region of the spike protein has been observed during prolonged infections in immunocompromised patients and proposed as a mechanism that evades the proofreading activity of the virus and accelerates adaptive evolution. The 141-144LGVY and 243-244LA deletions confer resistance to NTD specific monoclonal antibody in neutralization assays^15^. The Q493K/R and E484K substitutions are located in the RBD region of the spike protein, and associated with resistance to monoclonal antibodies or convalescent plasma^16,17^. In particular, the E484K mutation has been linked to the rapid spread of B.1.351 and B.1.1.28 variants in South Africa and Brazil, respectively. To date, over 17,000 E484K variants have been identified from >60 countries within various SARS-CoV2 lineages (www.gisaid.org) (**Fig2**), potentially posing significant challenges to vaccine efficacy and increased reinfection risk. Intriguingly, the co-occurrence of 141-144LGVY and E484K in the third tracheal aspirate specimen completely replaced other mutants, suggesting this haplotype may have compensated for a fitness cost or have higher antibody resistance level.

**Fig 2.**
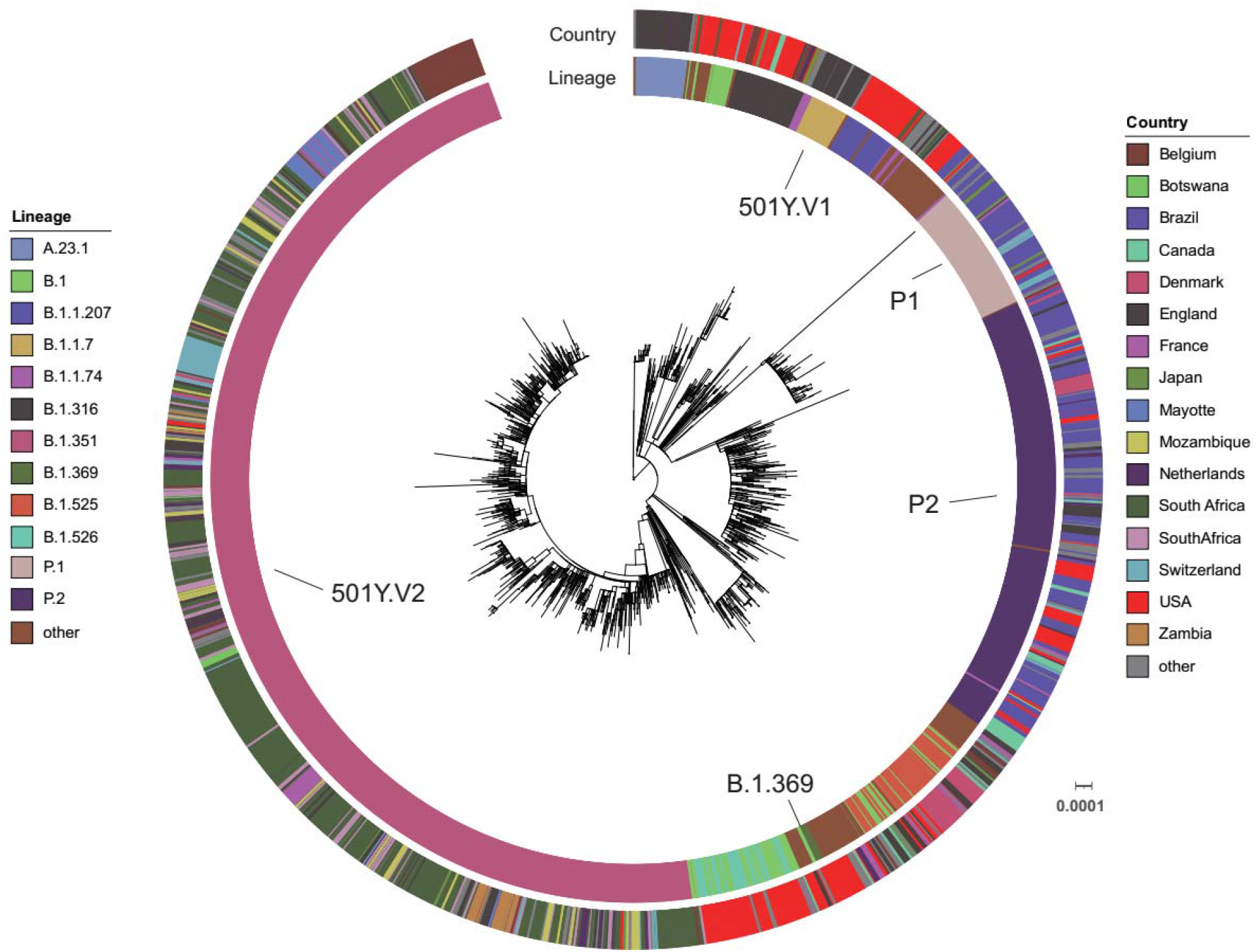
Global distribution of S protein E484K mutation. A maximum-likelihood phylogenetic tree with the 4 patient sequences (B.1.369) and 1,983 selected SARS-CoV-2 genomes from the GISAID database (date as 2/12/2021) is annotated using iTOL (www.itol.embl.de). The scale represents 0.0001 nucleotide substitutions per site. The SARS-CoV-2 Pangolin lineage and country are illustrated as two outer rings in different colors.

## Discussion

While most immunocompetent hosts are able to achieve resolution of COVID-19 within 1-3 weeks after symptoms, there is emerging evidence that a pre-existing immunocompromised state is associated with prolonged infection and significantly increased risk of severe disease^4,5,18,19^. Although the immunological mechanisms for control of SARS-CoV-2 in humans have not been clearly elucidated, it is likely that both cytotoxic T-cells and antibody-mediated immune responses are important for clearance of the viral infection^18,20^. Activation of type I and type III interferons has been postulated to be a key contributor to innate immune control ^20,21^, in addition to CD8^+^ effector T-cell–mediated killing of virally infected cells and CD4^+^ T cell– dependent enhancement of CD8^+^ and B-cell responses ^18,20^. Following viral clearance, long-term memory T-cells are formed for prolonged antiviral immunity. Chronic viral infection must evade or suppress some part of this pathway. Moreover, animal studies have shown that chronic viral infections were characterized by persistent antigenic activation of T-cells, ultimately driving a nonresponsive cell state known as T-cell “exhaustion”. This state was often accompanied by lymphopenia^20^. This patient’s anti-rejection regimen of mycophenolate and tacrolimus targets and inhibits T-cell function and replication, and treatment with prednisone may compromise the ‘proliferative burst’ of effector T-cells ^22^. While mycophenolate was discontinued, the patient was maintained on tacrolimus and prednisone during his entire hospitalization, which likely further impaired his cellular immunity against SARS-CoV-2. In our case, the treatment with convalescent plasma in combination with the routine maintenance of an anti-rejection regiment may have facilitated a “breeding ground” and the emergence of immune-escape mutants. Although we have no evidence that these two escape variants were transmitted, this case suggest that VOCs may arise among immunocompromised populations undergoing anti-SARS-CoV-2 therapy, and enhanced measures will be required to reduce transmission.

## Data Availability

All sequence data were deposited in the National. Center for Biotechnology Information database under. BioProject PRJNA675117.

## Acknowledgement

We gratefully acknowledge the Authors from the Originating laboratories responsible for obtaining the specimens and the Submitting laboratories where genetic sequence data were generated and shared via the GISAID Initiative, on which this research is based.

